# SARS-CoV-2 spillover into hospital outdoor environments

**DOI:** 10.1101/2020.05.12.20097105

**Authors:** Dayi Zhang, Yunfeng Yang, Xia Huang, Jingkun Jiang, Miao Li, Xian Zhang, Haibo Ling, Jing Li, Yi Liu, Guanghe Li, Weiwei Li, Chuan Yi, Ting Zhang, Yongzhong Jiang, Yan Xiong, Zhenyu He, Xinzi Wang, Songqiang Deng, Peng Zhao, Jiuhui Qu

## Abstract

The outbreak of coronavirus infectious disease-2019 (COVID-19) pandemic has rapidly spread throughout over 200 countries, posing a global threat to human health. Till 15^th^ May 2020, there are over 4.5 million confirmed cases, with roughly 300,000 death^1^. To date, most studies focus on severe acute respiratory syndrome coronavirus 2 (SARS-CoV-2) in indoor environment owing to its main transmission routes *via* human respiratory droplets and direct contact^2,3^. It remains unclear whether SARS-CoV-2 can spill over and impose transmission risks to outdoor environments despite potential threats to people and communities. Here, we investigated the presence of SARS-CoV-2 by measuring viral RNA in 73 samples from outdoor environment of three hospitals in Wuhan. We detected SARS-CoV-2 in soils (205-550 copies/g), aerosols (285-1,130 copies/m^3^) and wastewaters (255 to 18,744 copies/L) in locations close to hospital departments receiving COVID-19 patients or in wastewater treatment sectors. These findings reveal significant viral spillover in hospital outdoor environments that was possibly caused by respiratory droplets from patients or aerosolized particles from wastewater containing SARS-CoV-2. In contrast, SARS-CoV-2 was not detected in other areas or on surfaces with regular disinfection implemented. Soils may behave as viral warehouse through deposition and serve as a secondary source spreading SARS-CoV-2 for a prolonged time. For the first time, our findings demonstrate that there are high-risk areas in hospital outdoor environments to spread SARS-CoV-2, calling for sealing of wastewater treatment unit and complete sanitation to prevent COVID-19 transmission risks.

---

SARS-CoV-2 is an enveloped, positively-stranded RNA virus belonging to the beta coronavirus genus that causes COVID-19^4-6^. It can transmit among people ^5,7-9^ *via* direct contact and respiratory droplet routes^6,10,11^, while aerosol or faecal transmission route is also possible^12-14^. Many studies have analyzed SARS-CoV-2 in hospital indoor environment to assess its transmission dynamics and develop strategies to protect medical staffs or drop-in visitors^3,15^. In contrast, there is no knowledge about viral presence in outdoor environment. As a result, it is intractable to evaluate the potential spill-over into open space and distribution in outdoor environmental matrices of SARS-CoV-2, that potentially survives for a prolonged time and threatens public health.

We collected 73 outdoor environmental samples of water (28), aerosol (16), soil (15) and road- or wall-surface (14) from 13 sites in three specialized hospitals in Wuhan dedicated for COVID-19 treatments, i.e., Jinyintan Hospital, Huoshenshan Hospital and Wuchang Fangcang Hospital, during their operation receiving COVID-19 patients in March and April (Figure 1 and Table S1, see Methods for details). Three out of 15 soil samples (20%) exhibited positive results for SARS-CoV-2 viral RNA, which were collected near wastewater treatment sectors and outside patient departments (Table 1). Among them, the soil sample with 2-m distance to the adjusting tank in wastewater treatment sector of Jinyintan Hospital contained 253 copies/g of SARS-CoV-2 (Table S1), whereas the other sample 2-m away from the disinfection tank was negative in SARS-CoV-2. In other areas, there was 205 copies/g of SARS-CoV-2 in soil with 5 m distance to the outpatient department. In Huoshenshan Hospital, only soil with 2 m distance to the first adjusting tank contained SARS-CoV-2 viral RNA at a level of 550 copies/g, while soils in other wastewater treatment units or medical staff living area exhibited negative signals. None of soils in Wuchang Fang Hospital surrounding the septic tanks and in background areas was positive for virus. To our knowledge, this is the first report showing the presence of SARS-CoV-2 viral RNA in soils. Since soils are environmental matrices rich in organic matters that can protect and shield viruses, SARS-CoV-2 might not be sanitized by disinfectants or ultraviolet light as evidenced by previous studies on other viruses^16,17^, and possibly survive for a prolonged time in hospital outdoor environment.

**Figure 1.**
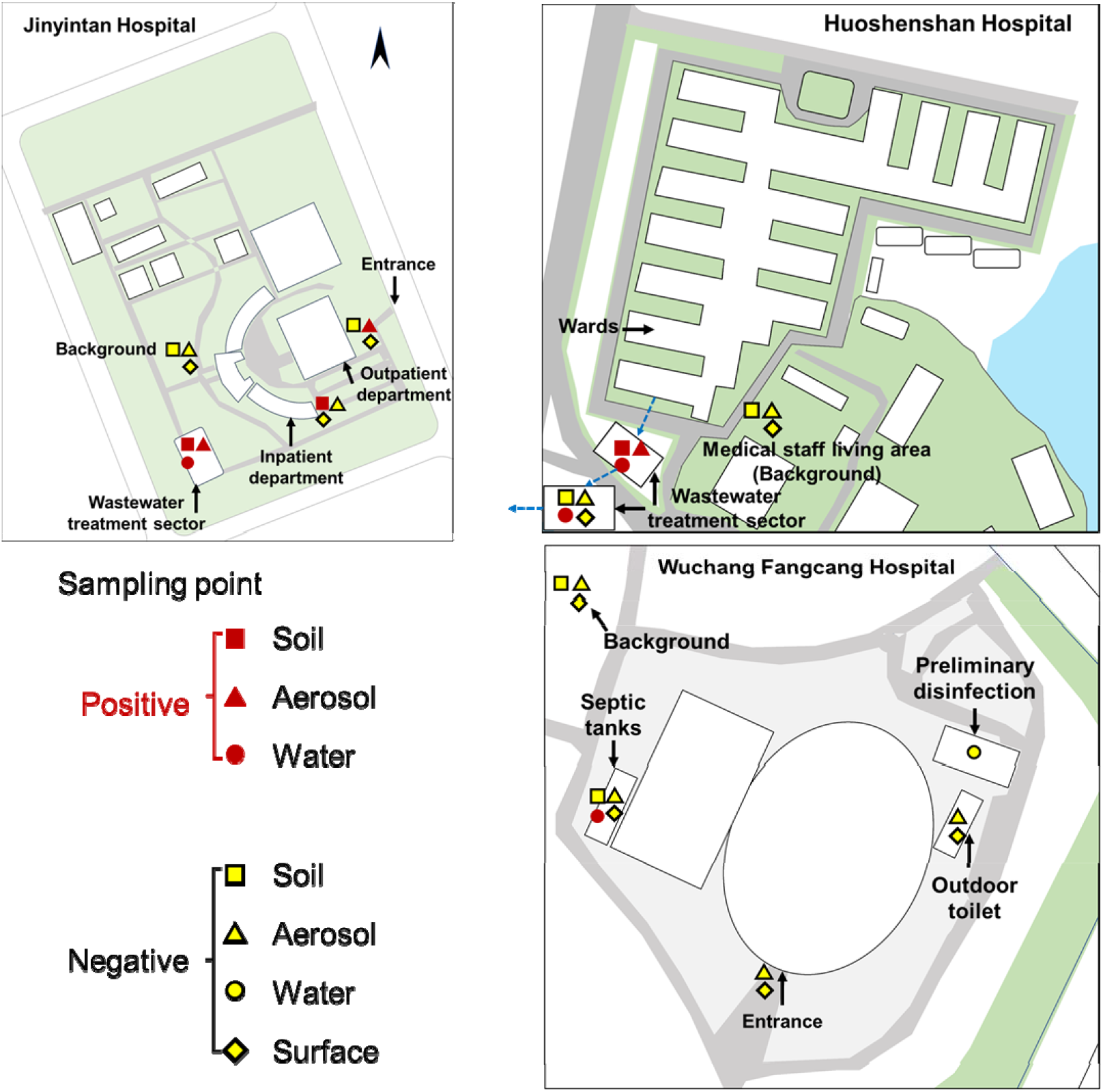
Outdoor environment sampling sites in Jinyintan Hospital, Huoshenshan Hospital and Wuchang Fangcang Hospital.

**Table 1.** Presence of SARS-CoV-2 in hospital outdoor environments.

| Hospital | Site | Sample type<sup>a</sup> |  |  |  |
| --- | --- | --- | --- | --- | --- |
|  |  | Soil<br>(copies/g) | Aerosol<br>(copies/m<sup>3</sup>) | Water<br>(copies/L) | Surface<br>(copies/m<sup>2</sup>) |
| Jinyintan | Wastewater treatment sector | ND<sup>b</sup>-253<br>(1/2) | 285<br>(1/1) | ND-255<br>(1/5) | - |
|  | Out- and In-patient department | ND-205<br>(1/2)) | ND-1,130<br>(1/2)) | - | ND<br>(0/2) |
|  | Background | ND<br>(0/1) | ND<br>(0/1) | - | ND<br>(0/1) |
| Huoshenshan | Wastewater treatment sector | ND-550<br>(1/2) | ND-603<br>(1/3) | ND-2,208<br>(3/10) | ND<br>(0/2) |
|  | Background (Medical staff living area) | ND<br>(0/2) | ND<br>(0/2) | - | ND<br>(0/2) |
| Wuchang<br>Fangcang | Wastewater treatment sector | ND<br>(0/4) | ND<br>(0/2) | ND-18,744<br>(7/13) | - |
|  | Entrance | - | ND<br>(0/2) | - | ND<br>(0/2) |
|  | Outdoor toilet | - | ND<br>(0/1) | - | ND<br>(0/2) |
|  | Background | ND<br>(0/2) | ND<br>(0/2) | - | ND<br>(0/2) |

<sup>a</sup> Fraction in bracket is number of positive samples to number of total samples.
<sup>b</sup> ND, non-detected.

All road- and wall-surfaces in outdoor environment of the three studied hospitals were negative in viral signals (Table 1). Surface contamination of SARS-CoV-2^18^, Middle East Respiratory Syndrome (MERS) coronavirus^19,20^ and norovirus^21^ has been observed in hospital indoor environment. Our results documented that frequent disinfection of three times a day in hospital outdoor environment effectively removed the virus of SARS-CoV-2.

To trace the source of SARS-CoV-2 in soils, we collected the surrounding aerosol samples. Inside the adjusting tank of Jinyintan Hospital and Huoshenshan Hospital, SARS-CoV-2 in aerosols was found at a level of 285 copies/m^3^ and 603 copies/m^3^, respectively (Table 1 and Table S1). They were of comparable levels to SARS-CoV-2 detected in intensive care units (ICU)^3,22^ and exhibited high transmission potential *via* aerosol deposition. On the contrary, aerosol SARS-CoV-2 was not detected in downstream wastewater treatment units of the second adjusting tank and the moving-bed biofilm reactor (MBBR). Outside patient departments of Jinyintan Hospital, SARS-CoV-2 in the aerosols collected 5 m away from outpatient building were 1130 copies/m^3^, whereas undetected in aerosols collected 5 m away from inpatient building. Since aerosols are highly dynamic, SARS-CoV-2 in aerosols depends on the sources of respiratory droplets or airborne viruses in a short period, which is largely attributed to the aerosolized droplets from the tidal breathing of COVID-19 patients^2^. In contrast, no SARS-CoV-2 was detected in aerosols above soils exhibiting negative results, *e.g*., background area of Jinyintan Hospital, medical staff living area of Huoshenshan Hospital, and the entrance and outdoor toilet of Wuchang Fangcang Hospital. Our results imply that hospitals receiving COVID-19 patients have high-risk outdoor areas (patient departments and wastewater treatment sector). SARS-CoV-2 contamination on ground surface was only previously reported in indoor studies, explained by deposition of airflow-displaced virus-laden droplets in COVID-19 patient living room^18^. The co-existence of SARS-CoV-2 in both soils and aerosols in the high-risk areas hints viral spillover, deposition and accumulation in soils from airborne SARS-CoV-2.

To further examine whether the aerosol and soil SARS-CoV-2 was derived from wastewater treatment sector, we analyzed SARS-CoV-2 viral RNA in waters from different treatment tanks. SARS-CoV-2 in Jinyintan Hospital was only detected in water from the adjusting tank (255 copies/L, Table S1), but undetected in other tanks and effluents. Raw medical wastewater in the adjusting tank of Huoshenshan Hospital contained 633 copies/L of SARS-CoV-2, which was only occasionally found in MBBR (505 copies/L) and sedimentation tank (2,208 copies/L) (Table S1). No SARS-CoV-2 was detected in effluents after disinfection. All water samples collected in the preliminary disinfection tanks of Wuchang Fangcang Hospital had no SARS-CoV-2 signal. SARS-CoV-2 RNA was detected in wastewater from the septic tanks disinfected by 800 mg/L of sodium hypochlorite, ranging from 557 to 18,744 copies/L, and it declined to non-detected after the dosage of sodium hypochlorite increased to 6,700 mg/L since 5^th^ March^23^. Our results suggest apparent presence of SARS-CoV-2 viral RNA in raw water from wards, signifying the need for complete disinfection. As it decayed rapidly in medical wastewater treatment process and a complete disinfection was applied for all effluents before discharge, there was negligible risk of SARS-CoV-2 spread through pipe network receiving treated wastewater from hospitals.

It is worthy of noting that SARS-CoV-2 was observed in all waters, aerosols and surrounding soils at the adjusting tank of wastewater treatment sector in Jinyintan Hospital and Huoshenshan Hospital (Table 1). As these areas are barely visited by COVID-19 patients and medical staffs, it is likely that SARS-CoV-2 arose from viral RNA-containing medical wastewater *via* aerosolization in the uplifting process, forming airborne virus-containing aerosols, and eventually depositing on soils. The wastewater treatment sector in Wuchang Fangcang Hospital is a temporary enclosed system effectively preventing the spillover of airborne SARS-CoV-2 from wastewater, resulting in negative viral signals in surrounding aerosols and soils. This implies appropriate sealing of the adjusting tank and other treatment units might block the potential viral transport from wastewater into aerosols and deposition on soils.

For the first time, we demonstrate the presence of SARS-CoV-2 viral RNA in hospital outdoor environments of three specialized hospitals dedicated for COVID-19 treatments. SARS-CoV-2 existed in all environmental matrices at hospital departments receiving confirmed or suspected COVID-19 patients (aerosols and soils) and wastewater treatment sector (waters, aerosols and soils), revealing high-risk areas for potential SARS-CoV-2 transmission as illustrated by Figure 2. High-risk areas located outside patient departments are exposed to respiratory droplets containing SARS-CoV-2 by receiving confirmed or asymptomatic COVID-19 patients. Alternatively, undisinfected medical wastewater in the adjusting tank of wastewater treatment sector might spill airborne viruses through uplifting or aeration and then deposit SARS-CoV-2 on surrounding soils and solid-surfaces. Traditional outdoor disinfection strategies mainly focus on walls, roads or facilities and can deactivate viruses on solid-surfaces with high efficiency^24,25^, explaining the negative results on all road- and wall-surfaces in this work. Viral presence and survival in soils are seldom examined and there is limited work addressing the potential risks of soil viruses^26^. Soils can receive the viruses from aerosols and waters, and potentially become a new source for SARS-CoV-2 transmission in outdoor environment. Non-point outdoor spillover of SARS-CoV-2 from individual houses of confirmed or asymptomatic COVID-19 patients and environmental viral residue in crowded areas need attentions and further investigations.

**Figure 2.**
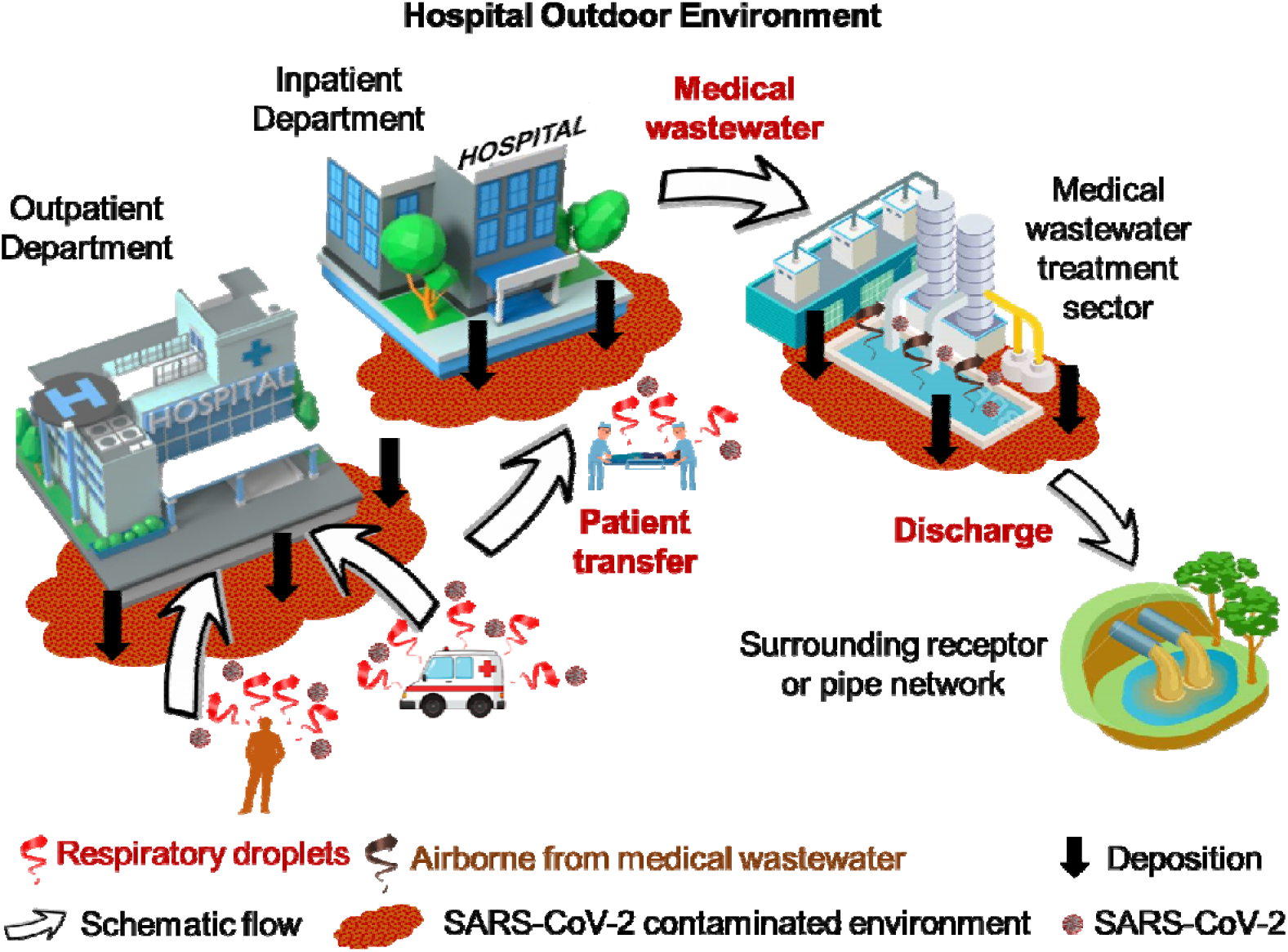
Spillover and potential transmission of SARS-CoV-2 in high-risk areas of hospital outdoor environments.

Although we do not address SARS-CoV-2 infectivity by viral culture and only collected limited numbers of samples owing to the strict control during the COVID-19 outbreak in Wuhan, China, our study unravels the distributions of SARS-CoV-2 in soils, aerosols, waters, and surfaces, covering the major outdoor environments of hospitals. Although viruses might decay rapidly from these high-risk areas and regular disinfection can effectively eliminate SARS-CoV-2, the overall risks of hospital outdoor environments are significant, particularly in those high-risk areas.

## Materials and methods

### Hospitals

Jinyintan Hospital is the first hospital in Wuhan receiving COVID-19 patients, and in outpatient and inpatient departments. Its medical wastewater treatment sector is comprised of an adjusting tank, an aeration tank for biodegradation, a secondary sedimentation tank and a disinfection tank (Figure S1A). Huoshenshan hospital is a newly-constructed hospital designated for COVID-19 patients and confirmed patents were transferred directly into wards. There is no inpatient or outpatient department, and the wastewater treatment sector consists of a process integrated storage sector with two adjusting tanks and one septic tank, a MBBR, a sedimentation tank and a disinfection tank (Figure S1B). Medical staff area is located south-east to ward area. Wuchang Fangcang Hospital is a temporary shielding hospital open from 5^th^ February to 10th March 2020, receiving 1124 COVID-19 patients. Wastewater from eight outdoor toilets were pumped in 4 preliminary disinfection tanks, transferred into three septic tanks outside, followed a final disinfection. After 24-hour, the effluent was pumped and discharged into pipe network and wastewater treatment plants. Chlorine-based disinfectants are supplemented in wards and the disinfection tank only.

### Sample collection

Sampling sites are located in outdoor environment of Jinyintan, Huoshenshan and Wuchang Fangcang Hospitals, including different in wastewater treatment sectors, inpatient departments, outpatient departments, outdoor toilets and temperate septic tanks (Figure 1 and Table S1). Around 2.0 L of water was directly collected in a plexiglass sampler. Aerosol samples were collected using bioaerosol samplers (WA-15, Beijing Dinglan Tech. Ltd., China) at a flow rate of 14.0 L/min for 30 min. About 20 grams of soils were sampled at the ground surface (< 5 cm), and surface samples were collected using swabs wetted with phosphate buffer saline (pH=7.4) from road or wall (20×20 cm). All samples were immediately placed in 4 °C ice-boxes and transferred into laboratory for RNA extraction on the same day.

### RNA extraction and RT-qPCR

RNA extraction from all environmental specimens followed our previously reported protocol ^23^ Briefly, water (400 mL) and aerosol (2-3 mL) samples were centrifuged at 3,000 rpm, and the supernatant was subsequently supplemented with NaCl (0.3 mol/L) and PEG-6000 (10%), settled overnight at 4 °C, and centrifuged at 10,000 g for 30 minutes. Soil (3-5 g) and viral pellets after PEG-6000 sedimentation was extracted using the RNeasy® PowerSoil® Total RNA Kit (MOBIO, Carlsbad, CA, USA) according to the manufacturer’s instructions. SARS-CoV-2 RNA was quantified by RT-qPCR using AgPath-ID™ One-Step RT-PCR Kit (Life Technologies, Carlsbad, CA, USA) on a LightCycler 480 Real-time PCR platform (Roche, Indianapolis, IN, USA) in duplicates. Two target genes simultaneously amplified were open reading frame lab (CCDC-ORF1, forwards primer:

5'-CCCTGTGGGTTTTACACTTAA-3'; reverse primer:

5'-ACGATTGTGCATCAGCTGA-3'; fluorescence probe:

5'-FAM-CCGTCTGCGGTATGTGGAAAGGTTATGG-BHQ1-3') and nucleocapsid protein (CCDC-N, forwards primer:

5'-GGGGAACTTCTCCTGCTAGAAT-3'; reverse primer:

5'-CAGACATTTTGCTCTCAAGCTG-3'; fluorescence probe:

5'-FAM-TTGCTGCTGCTTGACAGATT-TAMRA-3'). RT-qPCR amplification for CCDC-ORF1 and CCDC-N was performed in 25 μL reaction mixtures containing 12.5 μL of 2×RT-PCR Buffer, 1 μL of 25×RT-PCR Enzyme Mix, 4 μL mixtures of forward primer (400 nM), reverse primer (400 nM) and probe (120 nM), and 5 μL of template RNA. Reverse transcription was conducted at 45 °C for 10 min (1 cycle), followed by initial denaturation at 90 °C for 10 min (1 cycle) and 40 thermal cycles of 60 °C for 45 second and 90 °C for 15 seconds. Quantitative fluorescent signal for each sample was normalized by ROX™ passive reference dye provided in 2×RT-PCR buffer. For each RT-qPCR run, both positive and negative controls were included. The copy numbers of SARS-CoV-2 was obtained from a standard calibration curve by a 10-fold serial dilution of genes encoding nucleocapsid protein with an amplification efficiency of 102.6%, calculated as copies=10^[-(Cq-39086)/32621^ (R^2^=0.991). For quality control, a reagent blank and extraction blank were included for RNA extraction procedure and no contamination was observed.

## Data Availability

All data are available from the authors by request.

## Author Contribution

D.Z., G.L., Y.L. and J.Q. conceptualized the study design; D.Z., H.L., W.L. and C.Y. collected samples; D.Z., J.L., X.Z., T.Z., Y.J., Y.X., Z.H. and X.W. did the laboratory test; D.Z., Y.Y., X.H., J.J., M.L., G.L., S.D. and P.Z. interpreted the results; D.Z., Y.Y., J.J., X.H., M.L., S.D. and J.Q. drafted the manuscript. All authors read and approved the final manuscript.

## References

1 WHO. Coronavirus disease (COVID-19) Situation Report. 15 May, (2020).

2 Qian, H. et al. Indoor transmission of SARS-CoV-2. medRxiv, 2020.2004.2004.20053058, (2020).

3 Liu, Y. et al. Aerodynamic analysis of SARS-CoV-2 in two Wuhan hospitals. Nature, 10.1038/s41586–41020-42271-41583, (2020).

4 Ralph, R. et al. 2019-nCoV (Wuhan virus), a novel Coronavirus: human-to-human transmission, travel-related cases, and vaccine readiness. Journal of Infection in Developing Countries 14, 3–17, (2020).

5 Li, Q. et al. Early Transmission Dynamics in Wuhan, China, of Novel Coronavirus–Infected Pneumonia. New England Journal of Medicine 382, 1199–1207, (2020).

6 Lai, C.-C., Shih, T.-P., Ko, W.-C., Tang, H.-J. & Hsueh, P.-R. Severe acute respiratory syndrome coronavirus 2 (SARS-CoV-2) and coronavirus disease-2019 (COVID-19): The epidemic and the challenges. Int. J. Antimicrob. Agents 55, 105924, (2020).

7 Chan, J. F. W. et al. A familial cluster of pneumonia associated with the 2019 novel coronavirus indicating person-to-person transmission: a study of a family cluster. Lancet 395, 514–523, (2020).

8 Poon, L. L. M. & Peiris, M. Emergence of a novel human coronavirus threatening human health. Nat Med, 1-2, (2020).

9 Chang, D. et al. Epidemiologic and Clinical Characteristics of Novel Coronavirus Infections Involving 13 Patients Outside Wuhan, China. JAMA 323, 1092–1093, (2020).

10 Carlos, W. G., Dela Cruz, C. S., Cao, B., Pasnick, S. & Jamil, S. Novel Wuhan (2019-nCoV) Coronavirus. American Journal of Respiratory and Critical Care Medicine 201, P7-P8, (2020).

11 Wu, J. et al. Clinical Characteristics of Imported Cases of COVID-19 in Jiangsu Province: A Multicenter Descriptive Study. Clinical Infectious Diseases, ciaa199, (2020).

12 Holshue, M. L. et al. First Case of 2019 Novel Coronavirus in the United States. New England Journal of Medicine 382, 929–936, (2020).

13 Tian, Y, Rong, L., Nian, W. & He, Y. Review article: Gastrointestinal features in COVID-19 and the possibility of faecal transmission. Alimentary Pharmacology & Therapeutics, doi: 10.1111/apt.15731, (2020).

14 Zhang, J., Wang, S. & Xue, Y. Fecal specimen diagnosis 2019 Novel Coronavirus–Infected pneumonia. Journal of Medical Virology, https://doi.org/10.1002/jmv. 25742, (2020).

15 Chia, P. Y. et al. Detection of Air and Surface Contamination by Severe Acute Respiratory Syndrome Coronavirus 2 (SARS-CoV-2) in Hospital Rooms of Infected Patients. medRxiv, 2020.2003.2029.20046557, (2020).

16 Hurst, C. J., Gerba, C. P. & Cech, I. Effects of Environmental Variables and Soil Characteristics on Virus Survival in Soil. Appl. Environ. Microbiol. 40, 1067–1079, (1980).

17 Vettori, C., Gallori, E. & Stotzky, G. Clay minerals protect bacteriophage PBS1 of *Bacillus subtilis* against inactivation and loss of transducing ability by UV radiation. Canadian Journal of Microbiology 46, 770–773, (2000).

18 Ong, S. W. X. et al. Air, surface environmental, and personal protective equipment contamination by severe acute respiratory syndrome coronavirus 2 (SARS-CoV-2) from a symptomatic patient. JAMA 323, 1610–1612, (2020).

19 Kim, S.-H. et al. Extensive Viable Middle East Respiratory Syndrome (MERS) Coronavirus Contamination in Air and Surrounding Environment in MERS Isolation Wards. Clinical Infectious Diseases 63, 363–369, (2016).

20 Weber, D. J., Sickbert-Bennett, E. E., Kanamori, H. & Rutala, W. A. New and emerging infectious diseases (Ebola, Middle Eastern respiratory syndrome coronavirus, carbapenem-resistant Enterobacteriaceae, Candida auris): Focus on environmental survival and germicide susceptibility. American Journal Of Infection Control 47, A29-A38, (2019).

21 Morter, S. et al. Norovirus in the hospital setting: virus introduction and spread within the hospital environment. Journal Of Hospital Infection 77, 106–112, (2011).

22 Guo, Z.-D. et al. Aerosol and Surface Distribution of Severe Acute Respiratory Syndrome Coronavirus 2 in Hospital Wards, Wuhan, China, 2020. Emerging Infectious Diseases 26, 10.3201/eid2607.200885, (2020).

23 Zhang, D. et al. Potential spreading risks and disinfection challenges of medical wastewater by the presence of Severe Acute Respiratory Syndrome Coronavirus 2 (SARS-CoV-2) viral RNA in septic tanks of fangcang hospital. medRxiv, 2020.2004.2028.20083832, (2020).

24 Brady, M. T., Evans, J. & Cuartas, J. Survival and disinfection of parainfluenza viruses on environmental surfaces. American Journal Of Infection Control 18, 18–23, (1990).

25 Hota, B. Contamination, disinfection, and cross-colonization: Are hospital surfaces reservoirs for nosocomial infection? Clinical Infectious Diseases 39, 1182–1189, (2004).

26 Kuzyakov, Y. & Mason-Jones, K. Viruses in soil: Nano-scale undead drivers of microbial life, biogeochemical turnover and ecosystem functions. Soil Biol Biochem 127, 305–317, (2018).

